# Longitudinal Assessment of Glymphatic Changes Following Mild Traumatic Brain Injury: Insights from PVS burden and DTI-ALPS Imaging

**DOI:** 10.1101/2024.06.01.24307927

**Authors:** Jiachen Zhuo, Prashant Raghavan, Li Jiang, Steven Roys, Rosy Linda Njonkou Tchoquessi, Hegang Chen, Emerson M. Wickwire, Gunjan Y. Parikh, Gary T. Schwartzbauer, Lynn M. Grattan, Ze Wang, Rao P. Gullapalli, Neeraj Badjatia

## Abstract

Traumatic brain injury (TBI) even in the mild form may result in long-lasting post-concussion symptoms. TBI is also a known risk to late-life neurodegeneration. Recent studies suggest that dysfunction in the glymphatic system, responsible for clearing protein waste from the brain, may play a pivotal role in the development of dementia following TBI. Given the diverse nature of TBI, longitudinal investigations are essential to comprehending the dynamic changes in the glymphatic system and its implications for recovery. In this prospective study, we evaluated two promising glymphatic imaging markers, namely the enlarged perivascular space (ePVS) burden and Diffusion Tensor Imaging-based ALPS index, in 44 patients with mTBI at two early post-injury time points: approximately 14 days (14Day) and 6-12 months (6-12Mon) post-injury, while also examining their associations with post-concussion symptoms. Additionally, 37 controls, comprising both orthopedic patients and healthy individuals, were included for comparative analysis. Our key findings include: 1) White matter ePVS burden (WM- ePVS) and ALPS index exhibit significant correlations with age. 2) Elevated WM-ePVS burden in acute mTBI (14Day) is significantly linked to a higher number of post-concussion symptoms, particularly memory problems. 3) The increase in the ALPS index from acute (14Day) to the chronic (6-12Mon) phases in mTBI patients correlates with improvement in sleep measures. Furthermore, incorporating WM-ePVS burden and the ALPS index from acute phase enhances the prediction of chronic memory problems beyond socio-demographic and basic clinical information, highlighting their distinct roles in assessing glymphatic structure and activity. Early evaluation of glymphatic function could be crucial for understanding TBI recovery and developing targeted interventions to improve patient outcomes.

## Introduction

Traumatic brain injury (TBI) is a significant global health concern, affecting approximately 55 million people worldwide^1^. About 70% of all TBI is categorized as mild, with admission Glasgow Coma Scale (GCS) of 13-15^2^. However, even “mild” brain injuries may result in structural and functional changes in the brain, leading to long term post-concussion symptoms^3–7^. The most common post-concussion symptoms are cognitive in nature, such as memory loss, reduced information processing speed, problems with divided attention, and decline in executive functions^8–10^. TBI is also known as a risk factor in the development of neurodegenerative diseases later in life, such as Alzheimer’s Disease^11–14^.

While it is still unclear how TBI contributes to progressive neurodegeneration, increasing evidence points toward glymphatic dysfunction and impaired protein waste clearance as potential key factors in developing dementia^15, 16^. The glymphatic system, a structured fluid transport network, spans the periarterial and perivenous spaces (located in the perivascular space, PVS), as well as the brain’s interstitial spaces^17–19^. This system facilitates the exchange of solutes between cerebrospinal fluid (CSF) and the interstitial fluid, allowing waste products from the brain to be transported away. The glymphatic system is also shown to be most active during sleep, when the brain’s extracellular space expands, allowing CSF infiltration along the perivascular space ^20, 21^. Notably, experimental TBI has been shown to cause a 60% reduction in glymphatic flow, which persists for over a month^22^. Additional evidence from animal studies also supports the idea that glymphatic dysfunction contributes to subsequent neurodegeneration and cognitive decline^17, 23^.

Imaging the glymphatic system directly in humans poses significant challenges due to the necessity of intrathecally injecting gadolinium (Gd)-based contrast agents as CSF tracers to track their passage through the glymphatic system^24, 25^. However, this method is invasive and not suitable for widespread application in the general population. Several non-invasive glymphatic imaging techniques have been developed^26, 27^ as alternative approaches for assessing the glymphatic system. Among them, two most promising and broadly used methods are the structural MRI-based Enlarged PeriVascular Spaces (ePVS) method^28, 29^, and the Diffusion Tensor Imaging ALong the Perivascular Spaces (DTI-ALPS) method^30, 31^. Elevated ePVS burden, identified from T2-weighted MRI images or in junction with T1-weighted MRI images, could indicate blockage in interstitial fluid drainage^32^ and impaired glymphatic function^28, 33^. A high ePVS burden is linked to normal aging^34, 35^, TBI^36–38^, Alzheimer’s disease and related dementia^39, 40^, and sleep quality ^41, 42^. Recent research has shown correlations between increased ePVS burden and poor sleep and persistent post-concussion symptoms in TBI patients^36, 37, 43^. The DTI ALPS index, derived from diffusion tensor imaging (DTI), quantifies glymphatic diffusion activity by assessing diffusivity along the deep medullary veins at the level of the lateral ventricular bodies^30^. A higher ALPS index, indicative of increased glymphatic system activity, has been shown to correlate with better cognitive function and brain reserve in the aging population^44^.

Conversely, a lower ALPS index is linked to poor sleep^45, 46^ and an elevated risk of developing Alzheimer’s disease and related dementia ^30, 44, 47–49^. In TBI patients, cross-sectional studies have yielded mixed results, with some reporting a reduced ALPS index post-injury^50–52^ while others showing an increased index ^53^. This discrepancy may reflect the dynamic changes in white matter structure and glymphatic activity during TBI recovery.

Due to the heterogeneous nature of TBI and its association with various pre-existing risk factors impacting patient outcomes^54–56^, longitudinal studies are critical for understanding the dynamic alterations in the glymphatic system and gaining insights into its role in TBI recovery.

This study aims to investigate the longitudinal changes in glymphatic imaging markers, specifically ePVS burden and the DTI-ALPS index, in patients with mild TBI (mTBI) during the early post-injury phase and up to one year post-injury. We hypothesize that glymphatic imaging markers will correlate with patient symptom ratings, and that longitudinal changes in these markers will reflect underlying glymphatic alterations associated with TBI recovery. By elucidating these relationships, we hope to provide insights into the potential of glymphatic imaging markers as predictors of long-term outcomes in TBI patients.

## Material and Methods

### Participants

Participants in this study included mTBI patients, orthopedic control patients with no head injury, and healthy controls without mTBI or orthopedic injury. Age range for all participants was 18 – 60 year and included 58% male and 42% female. All participants were English speaking and had no contraindications for MRI. mTBI and orthopedic control patients were recruited from the R Adam Cowley Shock Trauma Center (STC) at the University of Maryland Medical Center. Inclusion criteria for the *<u>mTBI patients</u>* included (I1) an admission Glasgow Coma Score (GCS) of 13–15; (I2) a positive admission CT, or if negative CT, then with documented/reported loss of consciousness or amnesia and/or evidence of facial trauma; (I3) admission to the STC within 24hrs of injury. Exclusion criteria included (E1) penetrating head injury; (E2) status post trauma due to asphyxiation; (E3) history of prior head injury requiring medical attention within the last 5 years, or prior injury at any time which resulted in persistent neurological or psychological deficits; (E4) history of neurological and/or psychiatric illness such as brain tumor, multiple sclerosis, Alzheimer’s disease, bipolar disorder, schizophrenia, seizures, etc.; (E5) history of stroke or myocardial infarction; (E6) active duty military status; (E7) police custody or prisoner status; and (E8) pregnant women. For the *<u>orthopedic control group</u>*, the inclusion criteria were being admitted to the STC for injuries other than brain injury and negative head CT if available, along with criteria (I3) as previously stated for mTBI patients. The same exclusion criteria applied. *<u>Healthy controls</u>* were recruited through online advertisements and flyers posted on the University of Maryland Baltimore campus, or they may be family or friends of the recruited patients. The inclusion criteria included no brain abnormalities, along with all the same exclusion criteria. The study was approved by the Institutional Review Board of the University of Maryland Baltimore. All participants underwent informed consent and were compensated for their participation in the study.

A total of 63 mTBI patients, 22 orthopedic control patients (OC) and 20 healthy controls (HC) completed the MRI and behavioral assessments. For patients, their initial MRI visit was at approximately 14 days post their injury (14Day visit). mTBI patients also came back for follow-up visits at 6 months to 12 months (6-12Mon visit). All MRI data were manually inspected, and data with motion artifacts and missing data were excluded. Additionally, any clinically relevant incidental findings from MRI scans of the two control cohorts were identified and subsequently excluded from further analysis.

After data cleaning, a total of 44 mTBI patients were included who had both the 14Day visit and the 6-12Mon visit. A total of 37 controls, including 19 OCs and 18 HCs was also included. Controls had only one study visit. For all mTBI patients, their admission CT findings were obtained from electronic medical database. MRI imaging findings were based on radiology reads from the 14Day conventional MRI sequences (T1-weighted T2-weighted, FLAIR, SWI, and diffusion weighted images). Any findings from either the admission CT or 14Day MRI were considered as positive for “CT or MRI findings”.

### Behavioral Assessments

All socio-demographic information was based on participant self-report. Participants also completed self-reported symptoms on the Modified Rivermead Post-Concussion Symptoms Questionnaire (RPQ)^57^ and the Pittsburgh Sleep Quality Index (PSQI)^58^ at each study visit. RPQ covers 22 symptoms, each is rated through a 5-point ordinal scale: 0 = not experienced at all, 1 = no more of a problem, 2 = a mild problem, 3 = a moderate problem, and 4 = a severe problem. Note that while patients were directed to report symptoms arising after the brain injury, healthy controls were directed to report symptoms as they experienced in the past month to establish a prevalence of these symptoms in the general adult population. Patients reporting at least the mild symptom (RPQ rating ≥ 2) were considered symptomatic, similar as in the TRACK-TBI study^10^. Total RPQ scores were calculated, along with total number of symptoms (rated at least mild). The Pittsburgh Sleep Quality Index (PSQI) is a 19-item questionnaire that assesses the quality and patterns of sleep. This measure focuses on the participants’ usual sleep habits during the past month and has been previously used in studies of mTBI^37, 59, 60^.

### MRI Data Acquisition

All MRIs were acquired in a 3T Siemens Prisma scanner with a 64-channel head and neck coil. Imaging protocol included 3D T1-weighted MPRAGE, 3D T2-weighted SPACE, and diffusion-weighted MRI. Both T1w and T2w images were acquired with 1mm^3^ isotropic resolution, 256mm FOV, and 176 sagittal sections. Other imaging parameters for T1w were TE/TR/TI = 3.37ms/4000ms/1400ms, flip angle = 6L, BW = 200Hz/Px, iPAT factor of 2; and for T2w were TE/TR = 349ms/3200ms, Turbo factor 280, BW = 781 Hz/Px, iPAT factor of 2x2. Diffusion MRIs were acquired with a 3x simultaneous multi-slice (SMS) EPI sequence in 75 axial slices with FOV = 224mm, isotropic resolution of 2mm, TE/TR = 78ms/3500ms, 2 b=0s/mm^2^ volumes and 45 diffusion directions at b=1000s/mm^2^ and 90 diffusion directions at b=2500s/mm^2^. An additional B0 volume was acquired with reversed phase-encoding direction to correct for susceptibility artifacts.

### Image Process

#### Enlarged PVS (ePVS) Burden

We employed a previously described method for automatic segmentation of the ePVS burden using 3D T1w and T2w images^29, 43^. Briefly speaking, the 3D T1w and T2w images were first processed with the Human Connectome Project (HCP) pipeline^61^ for rigidly aligning both sets of images to the MNI space, up-sampled to 0.7mm resolution, followed with sub-cortical segmentation using the FreeSurfer software (version v6.0.1). Following intensity normalization based on white matter signal, enhanced perivascular space contrast (EPC) image was generated, followed with frangi filter to create a vesselness map. To create the final binary ePVS mask, a threshold ‘h’ of 0.0000002 combined with a minimum cluster size of 9 (∼3mm^3^) was used for white matter ePVS. A slightly lower h was used here than previously noted^29, 43^ as we found this combined with a minimum cluster size in our data provided the best balance between sensitivity in detecting ePVS and limiting false detection rate^43^. For the basal ganglia region, h was set at 0.0000003 with the same minimum cluster size.

ePVS volume was calculated from both the whole brain WM and BG regions based on the FreeSurfer segmented masks. To account for brain size differences, the ePVS burden^43^ was calculated as *ePVS burden = 100% x ePVS volume/region volume.* The corresponding region was either the whole brain white matter (WM-ePVS burden) or basal ganglia (BG-ePVS burden). **Figure 1** shows an example of the EPC images along with segmented ePVS masks on a healthy control and an mTBI patient of similar ages, for both the white matter regions and basal ganglia regions, along with calculated WM-ePVS burden and BG-ePVS burden. Notably due to the lower contrast between ePVS and gray matter in the basal ganglia regions, the BG-ePVS segmentation appeared to be less reliable than WM-ePVS, with some ePVS missing.

**Figure 1.**
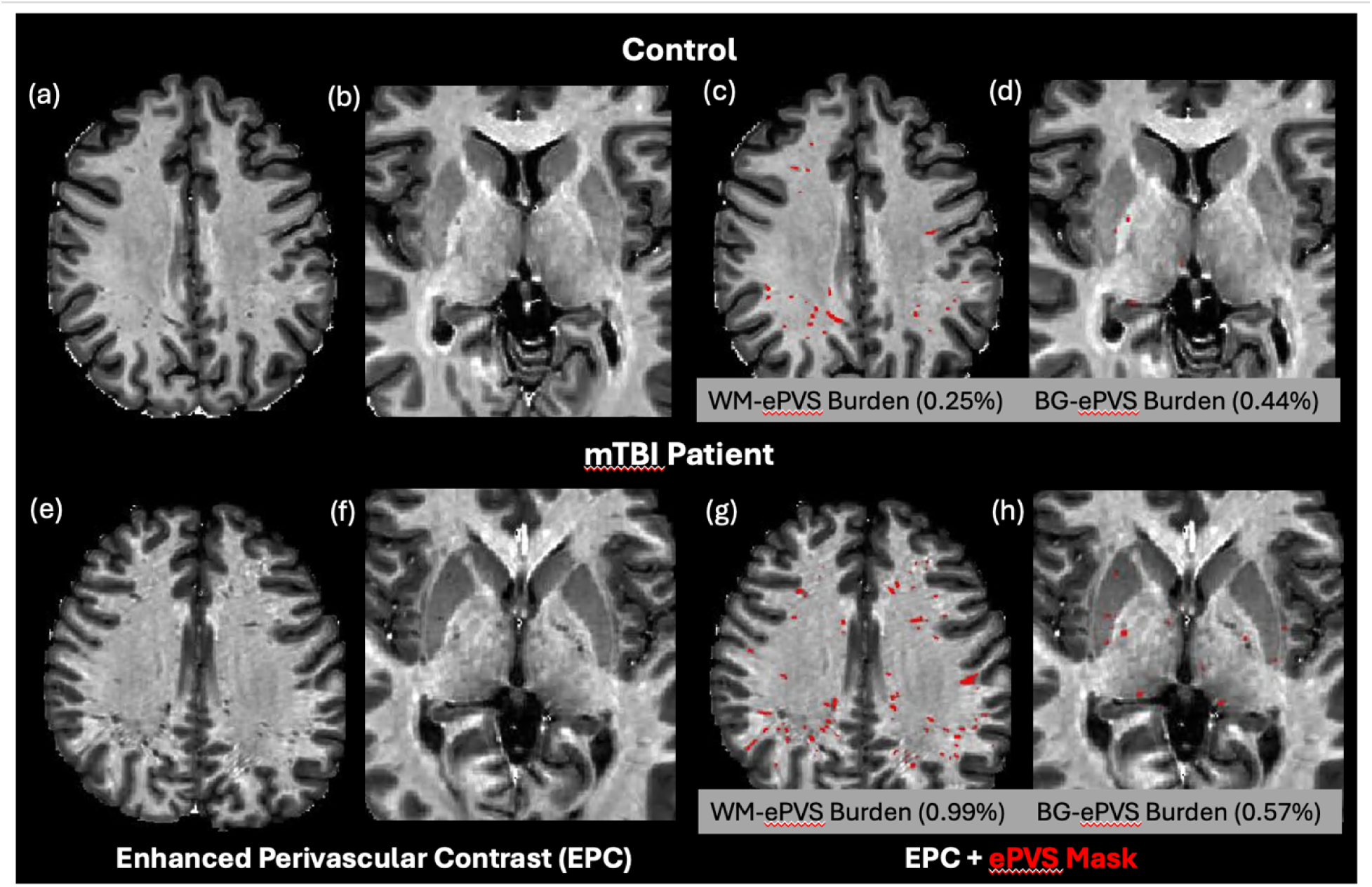
Enhanced Perivascular Contrast (EPC) images and segmented ePVS masks (in red) on a healthy control and TBI patients of similar age (20-30 years). Images (a, c, e, g) were ePVS segmentation in whole brain white matter (WM). Images (b, d, f, h) were ePVS segmentation in the basal ganglia (BG) region. Calculated WM-ePVS burden and the BG-ePVS burden were also shown for the corresponding cases.

### DTI-ALPS Index

DTI-ALPS index was calculated using the method originally proposed^30^. All diffusion-weighted images were preprocessed using the MRtrix3 software package^62^. Preprocessing steps included denoising^63^, eddy-current correction and motion correction^64^, and bias field correction using the N4 algorithm as provided in ANTs^65^.

Diffusion Kurtosis Imaging reconstruction were then performed using in-house Matlab program ^66, 67^ to estimate both a diffusion tensor and a kurtosis tensor. The diffusion tensor was then used to generate the diffusion coefficient maps along the X- (Dxx), Y-(Dyy), and Z- (Dzz) directions. For regions-of-interest (ROI) analysis, to improve reproducibility in ALPS calculation^68^, we took the approach to first warp all the diffusion maps to the MNI152 standard space provided in FSL (FMRIB Software Library, version 6.0, Oxford, UK)^69^. The B0 image was first registered to the HCP-processed T1w images using the EPI to structural registration tool, epi_reg (part of FSL). T1w images were warped to the MNI152 space using the SyN tool^70^ in ANTs. All registration steps were concatenated first before warping the diffusion maps to the MNI152 space.

Additionally, the principal diffusion direction map was warped using the vecreg tool (part of FDT-FMRIB’s Diffusion Toolbox) to create colored FA map in the MNI152 space.

Figure 2 shows the representative FA colormap and diffusivity maps on a representative participant, where four 16mm^2^ square ROIs were manually drawn in the projection fiber and association fiber adjacent to the medullary veins at the level of the lateral ventricle body. Placement of the ROIs were checked on all subjects and manually adjusted (if needed) to ensure proper coverage within the projection and association fiber regions. Mean values from the two bi-lateral ROIs were extracted for ALPS index calculation.

**Figure 2.**
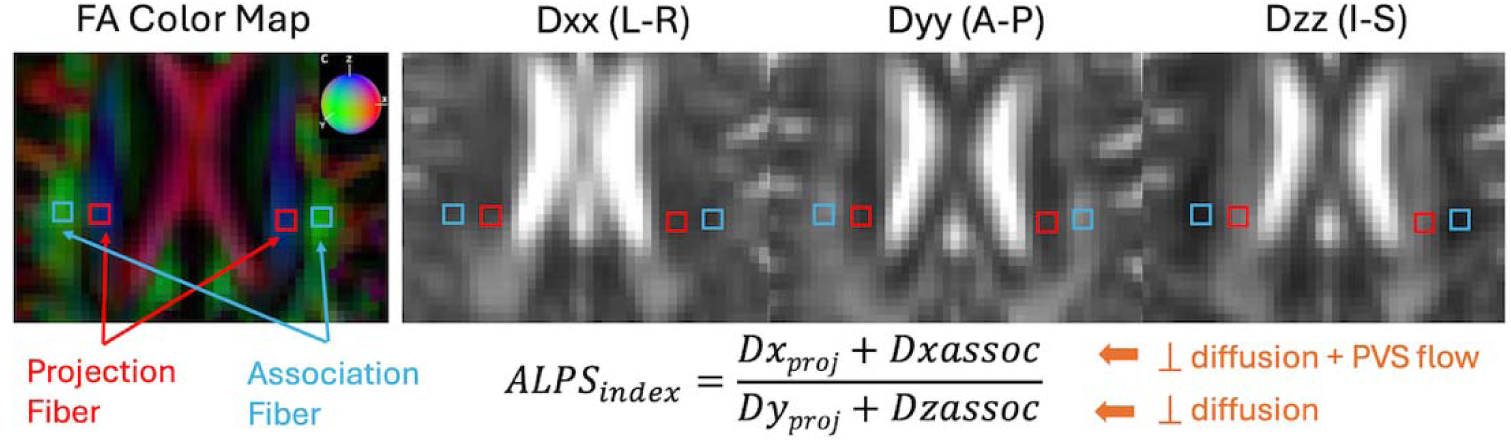
Representation and calculation of the DTI-ALPS index are shown on an example subject from the projection fiber (Dx_proj_ and Dy_proj_) and the association fiber (Dx_assoc_ and Dz_assoc_). Dxx, Dyy, and Dzz are diffusion coefficient maps along the X-, Y-,and Z-directions. The color sphere in the FA color map indicates the color coding of the diffusion tensor. L-R: Left-Right, A-P: Anterior-Posterior, I-S: Inferior-Superior.

### Data Analysis

Glymphatic imaging markers included WM-ePVS burden, BG-ePVS burden and ALPS index. Participant socio-demographic information included age, sex, years of education. Patient clinical information included admission GCS, binary indicator of positive CT or MRI findings (based on admission CT or 14Day-MRI). Participant assessment scores included total RPQ score for symptoms (range: 0 – 88), individual symptom score (0 – 4), total number of symptoms (0 – 22), and PSQI scores for sleep quality (0 – 21).

We first tested for differences of imaging markers and symptom scores between the two control cohorts (OCs and HCs) using two-sample t-tests and chi-squared tests. If no significant differences were found, we combined the two control cohorts into a single control group. We sub-grouped mTBI patients into those with and without memory problems based on their 14Day memory symptoms (RPQ rating ≥ 2). Group comparisons (controls vs. mTBI, mTBI- Memory Prob vs. mTBI-No Memory Prob) were conducted using two-sample t-tests for continuous variables and chi-squared tests for categorical variables.

Associations between imaging markers and age were analyzed using Pearson correlations. Associations of imaging measures with assessment scores were analyzed using multiple variable linear regression with age and sex as covariables. Longitudinal analysis in mTBI patients was conducted using linear mixed model with visit (14Day, 6-12Mon) as fixed effect, patient as a random effect, and age and sex as covariates. The ratio of imaging markers and assessment scores (6-12Mon / 14Day) were calculated to examine longitudinal changes over time.

Logistic regression with stepwise selection was used to develop prediction models for binary memory problems (RPQ rating ≥ 2) at the 6-12Mon visit, including socio-demographic (Age, Sex, Education), clinical information (GCS, CT or MRI findings), and glymphatic imaging markers (WM-ePVS, BG-ePVS, ALPS index) at the 14Day visit as independent variables. Receiver Operating Characteristic (ROC) curves were generated to compare the basic model (socio-demographic and clinical information) with the model including 14Day glymphatic imaging markers. Model comparisons were visualized using ROC curves, and area under the curves (AUC) were compared using the bootstrap method.

All statistical analyses were performed in R version 4.3.0 (R Core Team, 2021). Multiple comparisons were corrected by the False Discovery Rate (FDR) method. The significance level was defined as p < 0.05.

## Results

### Orthopedic Control (OC) vs. Healthy Control (HC)

Between-groups differences in socio-demographic, symptom ratings and glymphatic imaging markers between the orthopedic control and the healthy controls are presented in **Table 1**. There were no significant differences in age, any symptom ratings, or any glymphatic imaging markers. HCs did include more females (p = 0.043) and had significantly higher education years (p = 0.0003).

**Table 1.**
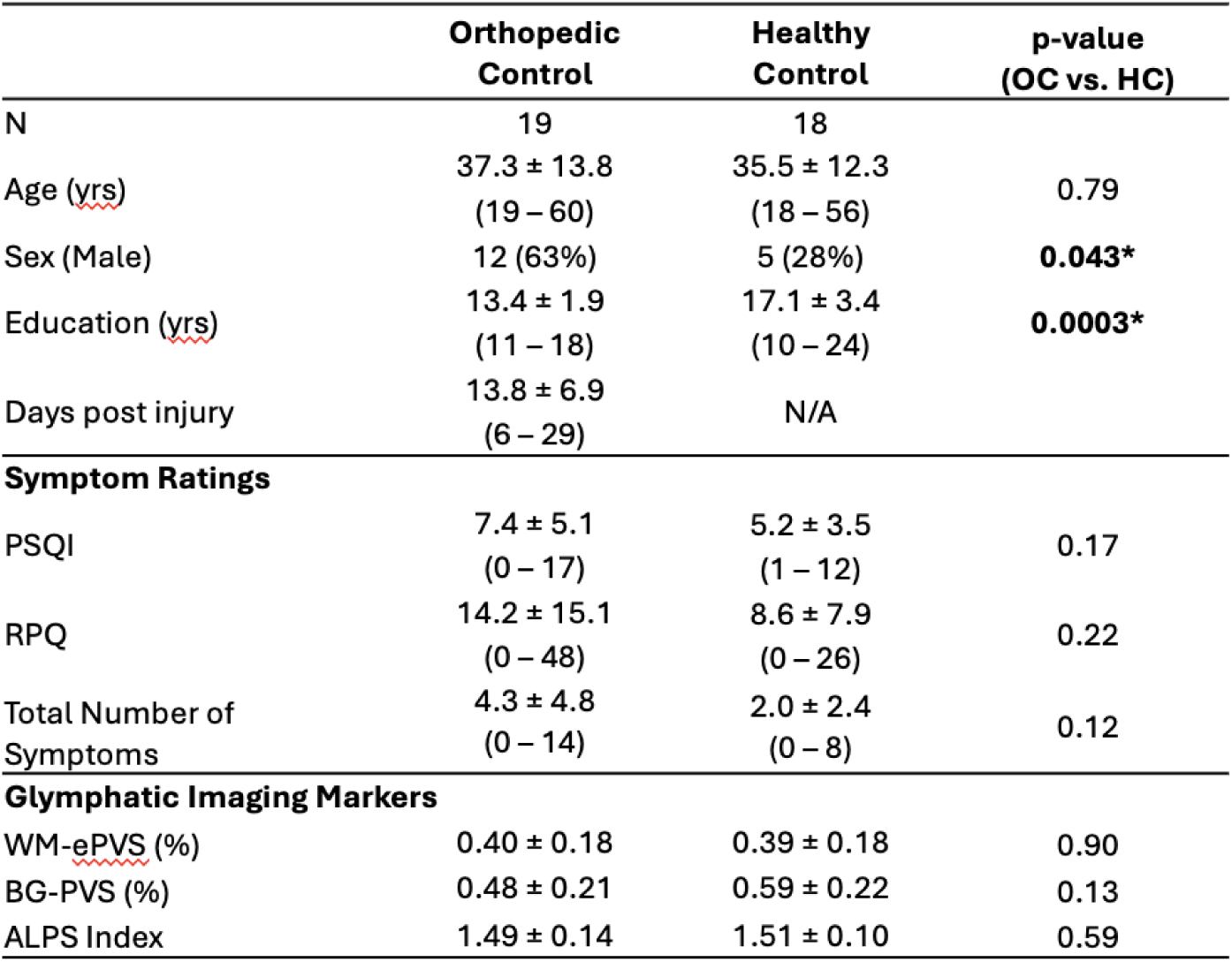
Socio-demographic, symptoms, and imaging measures for the two control groups (orthopedic control, OC, and healthy controls, HC). Values shown are Mean ± SD, with ranges or percentage in (). PSQI is the global PSQI score using the Pittsburgh Sleep Quality Questionnaire. RPQ is the total scores from the Rivermead Post Concussion Symptoms questionnaire. Total Number of Symptoms was counted when reported as at least mild symptoms (RPQ ratings ≥ 2). WM-ePVS is the white matter ePVS burden. BG-PVS is the basal ganglia ePVS burden. Significant p-values (p < 0.05) were shown in bold with *.

When combined to a single control cohort, the combined sex and education level did not significantly differ than the mTBI cohort (**Table 2**). We therefore combined the OCs and HCs to increase the sample size for controls.

**Table 2.**
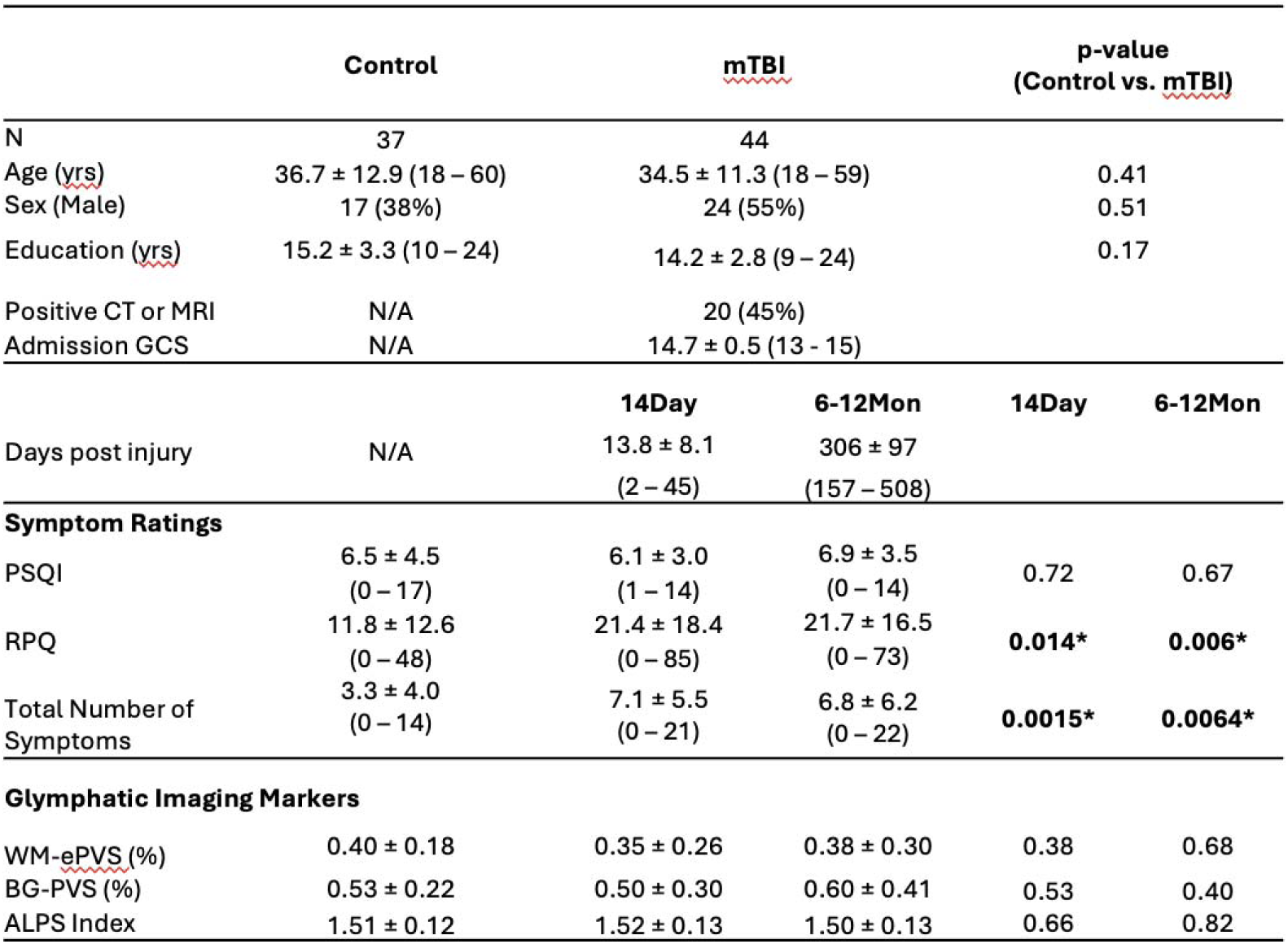
Socio-demographic, symptoms, and imaging measures for control and mTBI patients at two visits (14Day and 6-12Mon). Values shown are Mean ± SD, with ranges or percentage in (). Positive CT or MRI findings are based on admission CT and clinical MRI sequences. PSQI is the global PSQI score using the Pittsburgh Sleep Quality Questionnaire. RPQ is the total scores from the Rivermead Post Concussion Symptoms questionnaire. Total Number of Symptoms was counted when reported as at least mild symptoms (RPQ ratings ≥ 2). WM-ePVS is the white matter ePVS burden. BG-PVS is the basal ganglia ePVS burden. Significant p-values (p < 0.05) were shown in bold with *.

### Control vs. mTBI (14Day and 6-12Mon)

There were no differences in age, sex, education years between the mTBI patients and the controls. 45% of mTBI patients had either a positive finding from their admission CT or Day14 MRI. The mean days-post-injury for the two visits were 13.8 days (Day14) and 306 days (6-12Mon).

Interestingly, mTBI patients had similar PSQI scores as the controls at both visits, indicating that sleep problems may be a prevalent issue even in the general adult population. mTBI patients had significantly higher RPQ scores (Day14: p = 0.014; 6- 12Mon: p = 0.006) and total number of symptoms (Day14: p = 0.0015; 6-12Mon: p = 0.0064) than the controls, as expected.

The following symptoms were found to persist in mTBI patients at the 6-12Mon visit: sleep (54%), memory problems (44%), slow thinking (41%), trouble concentrating (59%), anxiety (51%), depression (46%), and fatigue (44%), where more than 40% of patients were still reporting at least a mild problem (RPQ rating ≥ 2). Figure 3 shows incidence of these symptoms (at least mild) for all groups at 6-12Mon. A significantly higher percentage of mTBI patients reported problems with cognitive symptoms, such as memory problems (p < 0.005), slow thinking (p < 0.005), and trouble concentrating (p < 0.005), but no differences were observed in other symptoms (sleep, anxiety, depression, or fatigue).

**Figure 3.**
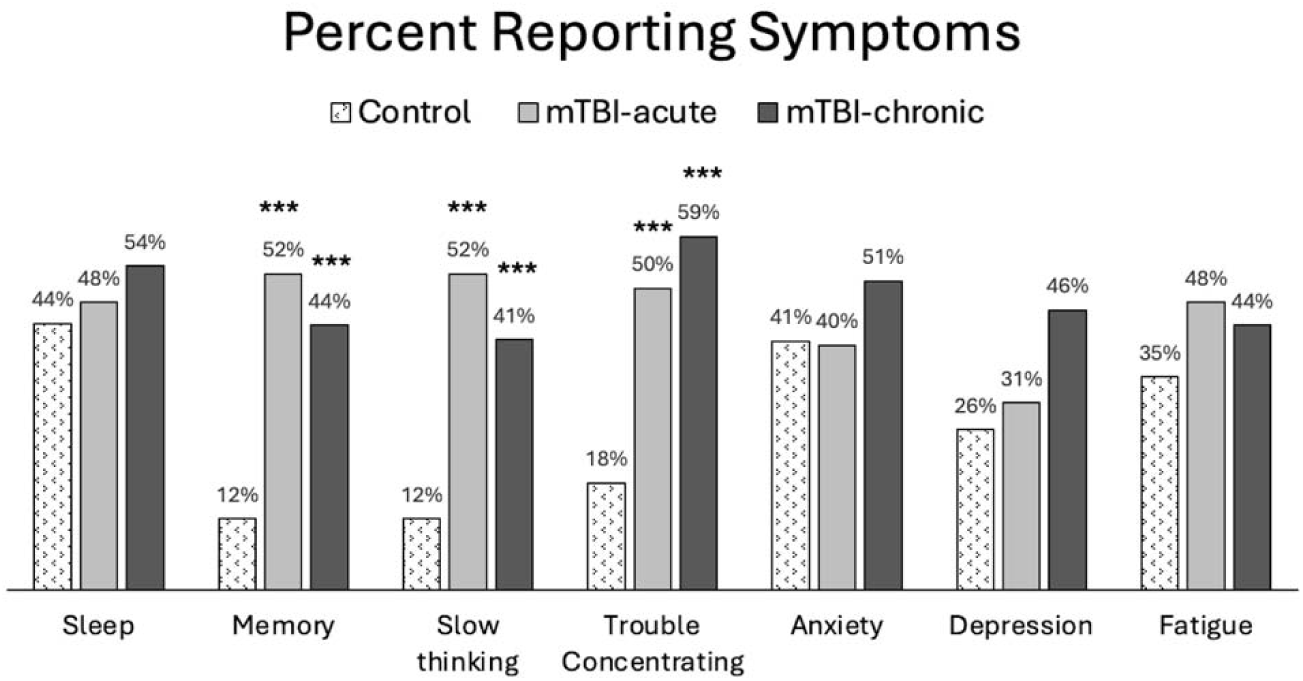
Percent population reporting at least mild symptoms (RPQ ratings ≥ 2) in controls and mTBI patients. Group comparisons were made between mTBI patients and controls using chi-square tests. ***: p < 0.0005.

No significant differences were observed in the glymphatic imaging markers between the controls and mTBI patients at either the Day14 or the 6-12Mon visit.

### Effect of Age of glymphatic imaging markers

Both WM-ePVS burden and ALPS index were found to be significantly correlated with age for both controls and mTBI patients. Figure 4 shows the scattered plots of the ALPS index and WM-ePVS burden with age in control and mTBI patients at both visits. In all groups, ALPS index significantly decreases with age (Control: r = -0.37, p = 0.024; mTBI 14Day: r = -0.40, p = 0.0071; mTBI 6-12Mon: r = -0.42, p = 0.0045) while the WM-ePVS burden significantly increases with age (Control: r = 0.51, p = 0.0013; mTBI 14Day: r = 0.45, p = 0.0022; mTBI 6-12Mon: r = 0.38, p = 0.011). The age-slope is however not significantly different in mTBI patients at each visit as compared to controls. Noticeably the variability of imaging markers is higher in mTBI patients, especially in WM-ePVS burden where more outlier high WM-ePVS were spotted in mTBI patients. One of the outlier cases (mTBI 14Day, WM-ePVS burden = 0.99%) was represented in Figure 1. No significant age-correlations were found for BG-ePVS burden (Control: p = 0.53, mTBI Day14: p = 0.77, mTBI 6-12Mon: p = 0.12).

**Figure 4.**
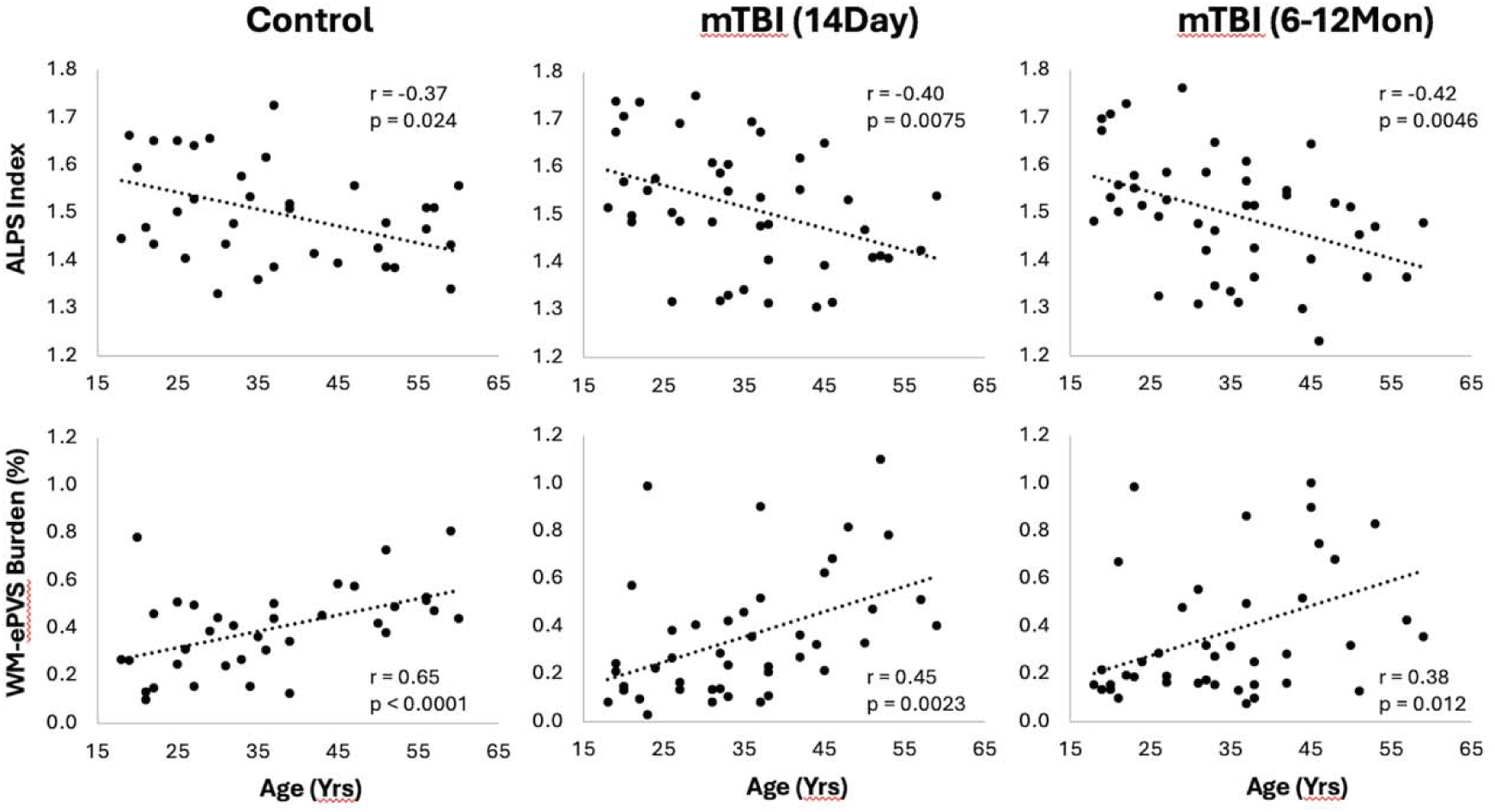
Scatter plots of ALPS index and WM-ePVS burden with age in controls and mTBI patients at both visits. Pearson correlation coefficient, r, and p-value were also shown for each group.

### Correlations between the glymphatic imaging markers

We also tested correlations between the glymphatic imaging markers among themselves for controls and mTBI at both visits. WM-ePVS burden was found to be positively correlated with the BG-ePVS burden (r = 0.34, p = 0.044), and negatively correlated with ALPS index (r = -0.36, p = 0.028). However, no significant correlations were found between these imaging markers in mTBI patients at any visit.

### Association of glymphatic imaging markers with symptoms

No significant correlations were found between glymphatic imaging markers with any symptom ratings in controls, or mTBI patients at the 6-12Mon visit.

For mTBI patients at the 14Day visit, the WM-ePVS burden were significantly correlated with the total number of symptoms (p = 0.0023) (Figure 5a), and more specifically memory problems (p = 0.017), based on RPQ ratings of ≥ 2 (at least with mild symptoms). We further divided mTBI patients based on their memory problems at the 14Day visit (RPQ memory ratings of ≥ 2), to two sub-groups: “No Memory Prob” and “With Memory Prob” (**Table 3, Figure5b**). The two patient sub-groups had no significant differences in age, sex, education, days post injury, or the PSQI scores. Patients with memory problems do present with significantly higher RPQ score (p < 0.0001) and symptom number (p < 0.0001). The WM-ePVS burden was significantly higher in the “With Memory Prob” group than the “No Memory Prob” group (p = 0.016). ALPS indexes do not differ between the two groups. Following up to the 6-12Mon visit, these two mTBI patient sub-groups continue to have significantly different WM-ePVS burden (p = 0.014), even though 10 patients had improving memory symptoms, while another 7 had worsening memory symptoms. The “With Memory Prob” group also continue to have significantly higher RPQ score (p = 0.02) and symptom number (p = 0.004), although the differences were smaller, likely indicating a recovery. No differences in ALPS index were observed.

**Figure 5.**
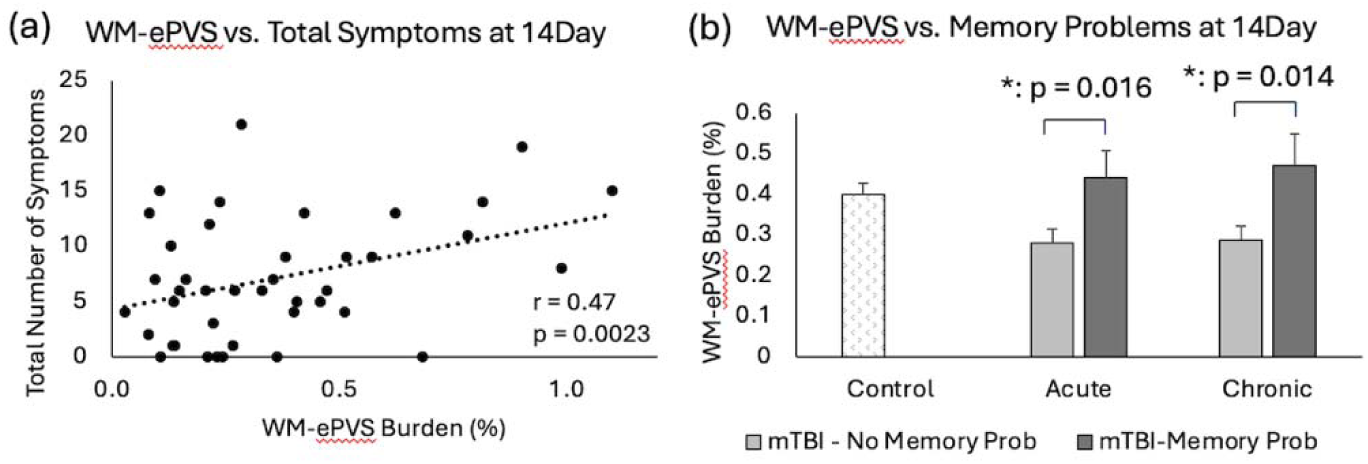
Associations of WM-ePVS burden with symptoms at 14Day in mTBI patients. (a) Scatter plots of WM-ePVS Burden and the total numbers of symptoms (with RPQ ratings ≥ 2) at 14Day, with age and sex-adjusted partial correlation coefficient and p-value. (b) WM-ePVS Burden in control and mTBI patients grouped by their reported memory problems (RPQ memory rating ≥ 2) at the 14Day visit.

**Table 3.** Sicio-demographic, symptoms, and imaging measures for mTBI patients with and without memory problems at two visits (14Day and 6-12Mon). The existence of memory problems were defined as RPQ ratings ≥ 2 at the 14Day visit. Values shown are Mean ± SD, with ranges or percentage in (). Positive CT or MRI findings are based on admission CT and clinical MRI sequences. PSQI is the global PSQI score using the Pittsburgh Sleep Quality Questionnaire. RPQ is the total scores from the Rivermead Post Concussion Symptoms questionnaire. Total Number of Symptoms was counted when reported as at least mild symptoms (RPQ ratings ≥ 2). WM-ePVS is the white matter ePVS burden. BG-PVS is the basal ganglia ePVS burden. Same mTBI patients were included for both visits, therefore demographic information stays the same. Significant p-values (p < 0.05) were shown in bold with *.

|  | No Memory Prob at 14Day |  | With Memory Prob at 14Day |  | Group Difference (p-value) |  |
| --- | --- | --- | --- | --- | --- | --- |
| N | 21 |  | 23 |  |  |  |
| Age (yrs) | 35.3 ± 11.2 (19 – 59) |  | 33.9 ± 11.5 (18 – 53) |  | 0.68 |  |
| Sex (Male) | 13 (62%) |  | 11 (48%) |  | 0.35 |  |
| Education (yrs) | 14.4 ± 3.2 (9 – 24) |  | 14.1 ± 2.4 (9 – 18) |  | 0.74 |  |
| Positive CT or MRI | 13 (62%) |  | 7 (30%) |  | 0.067 |  |
| Admission GCS | 14.7 ± 0.7 (13 – 15) |  | 14.9 ± 0.4 (14 – 15) |  | 0.25 |  |
|  | **14Day** | **6-12Mon** | **14Day** | **6-12Mon** | **14Day** | **6-12Mon** |
| Days post injury | 12.8 ± 9.1<br>(2 – 45) | 14.6 ± 7.1<br>(3 – 34) | 306 ± 99<br>(157 – 440) | 305 ± 97<br>(173 – 508) | 0.47 | 0.95 |
| **Symptom Ratings** |  |  |  |  |  |  |
| PSQI | 5.7 ± 3.2<br>(1 – 14) | 6.7 ± 4.0<br>(0 – 14) | 6.5 ± 2.8<br>(3 – 13) | 6.5 ± 2.8<br>(3 – 14) | 0.42 | 0.72 |
| RPQ | 7.8 ± 7.1<br>(0 – 21) | 15.1 ± 15.6<br>(0 – 46) | 33.8 ± 16.7<br>(15 – 85) | 27.2 ± 15.4<br>(4 – 73) | **< 0.0001\*** | **0.02\*** |
| Total Number of Symptoms | 2.7 ± 2.5<br>(0 – 7) | 4.0 ± 4.8<br>(0 – 15) | 11.0 ± 4.3<br>(6 – 21) | 9.3 ± 6.3<br>(1 – 22) | **< 0.0001\*** | **0.004\*** |
| **Glymphatic Imaging Markers** |  |  |  |  |  |  |
| WM-ePVS (%) | 0.27 ± 0.16 | 0.28 ± 0.17 | 0.44 ± 0.31 | 0.47 ± 0.36 | **0.016\*** | **0.014\*** |
| BG-PVS (%) | 0.43 ± 0.27 | 0.53 ± 0.32 | 0.56 ± 0.32 | 0.76 ± 0.64 | 0.17 | 0.14 |
| ALPS Index | 1.53 ± 0.13 | 1.49 ± 0.14 | 1.50 ± 0.13 | 1.50 ± 0.12 | 0.44 | 0.82 |

### Longitudinal Changes

No significant longitudinal differences were observed in mTBI patients from 14Day to 6-12Mon in PSQI, RPQ score or symptom numbers. The BG-ePVS burden significantly increased over time (p = 0.013), while other imaging markers were unchanged.

When investigating the visit effect and symptom ratings for imaging markers, the ALPS index demonstrated a significant positive association with the global PSQI score (p = 0.0062). Furthermore, the ratios of the ALPS index and the PSQI score (6- 12Mon/14Day) were also significantly correlated (r = -0.48, p = 0.008) (Figure 6).

**Figure 6.**
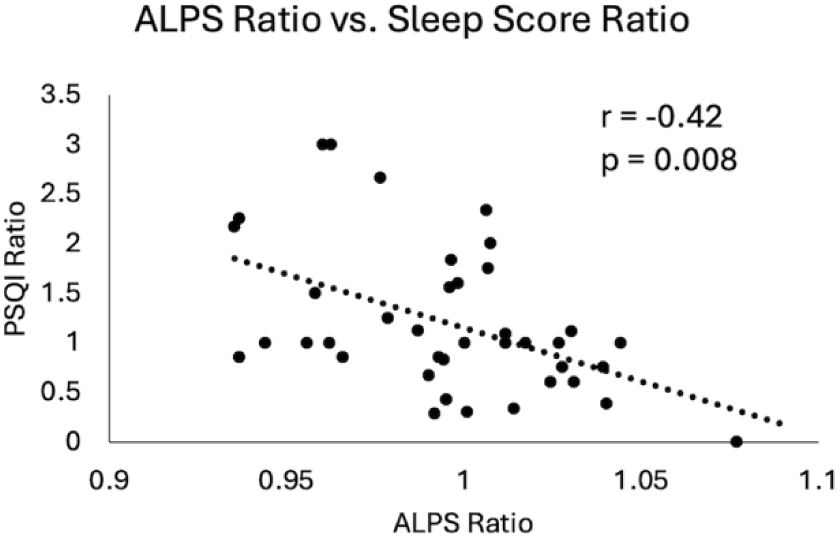
Scatter plot of changes in ALPS index and changes in sleep scores in mTBI patients from 14Day to 6-12Mon. Both the ALPS ratio and the PSQI ratio were calculated by dividing the 6-12Mon values by the 14Day values. Pearson correlation coefficient and p-value were displayed here.

Higher ALPS ratio, indicative of increase in the ALPS index from 14Day to 6-12Mon (increase glymphatic activity), was associated with reduction in PSQI scores (improvement in sleep condition).

### Prediction of 6-12Mon memory problem

In the mTBI patient cohort, we explored two models to predict their 6-12Mon memory outcome, based on existence of their memory problems (RPQ memory rating ≥ 2). The two models were <u>Model 1</u>: only with patients’ socio-demographic (Age, Sex, Education) and clinical information at 14Day (admission GCS, and CT or MRI findings). This represents the scenario for patients with basic clinical information and admission imaging findings. <u>Model 2</u> included all parameters in Model 1 and the glymphatic imaging markers at 14Day (WM-ePVS, BG-ePVS and ALPS). Stepwise selection selected two parameters for <u>Model 1</u>: Education and CT or MRI findings; and four parameters for <u>Model 2</u>: Education, CT or MRI findings, WM-ePVS and ALPS. The coefficients for Model 2 are shown in **Table 4**. The ROC curve for Model 2 as compared to Model 1 was shown in Figure 7, with AUCs of 0.68 for Model 1 and 0.84 for Model 2. Adding WM-ePVS and ALPS measured at 14Day in Model 2 significantly improved model prediction for 6-12Mon memory problems (p = 0.044).

**Figure 7.**
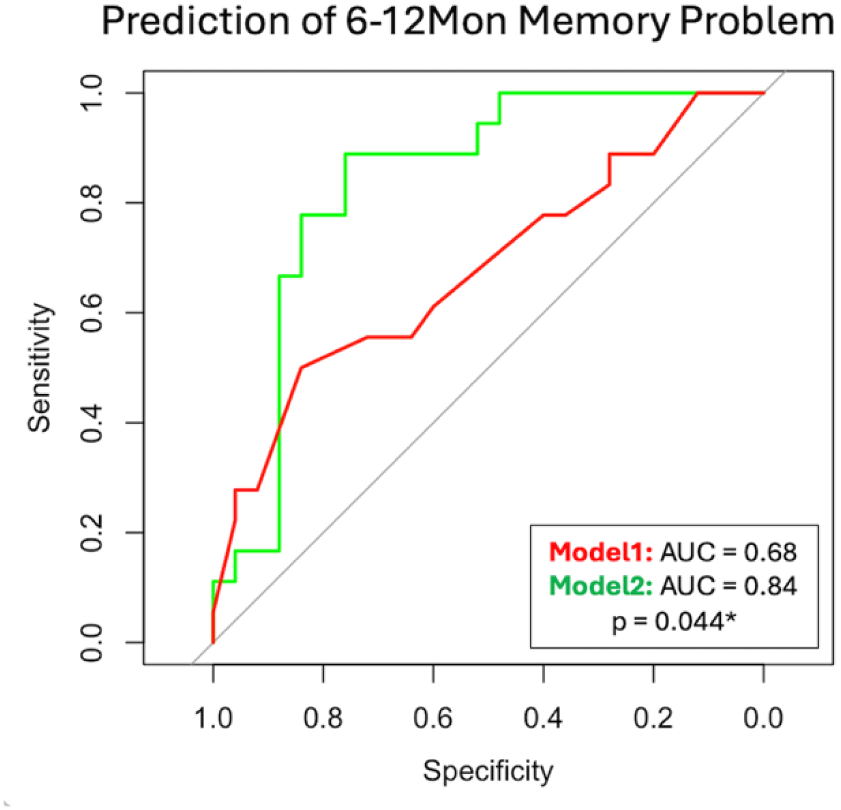
ROC curve for two logistic regression models predicting patient memory problems at the 6-12Mon visit (defined as RPQ rating ≥ 2). Model 1 used socio- demographic and clinical information, while Model 2 added 14Day glymphatic imaging measures. Both models were developed using step-wise selection. Model 1 included Education and CT or MRI findings, whereas Model 2 included Education, CT or MRI findings, WM-ePVS, and ALPS. The models were compared using the bootstrap method.

**Table 4.** Logistic regression model coefficients for Model 2 (Figure 7). Among all socio- demographic and clinical information, Education and CT or MRI findings were selected to compose the best logistic model for prediction of 6-12Mon memory problem. 14Day WM-ePVS and ALPS were also significant factors (bolded with *).

| Coefficients | Estimate Std. | Error | Z value | Pr(>\|z\|) |
| --- | --- | --- | --- | --- |
| (Intercept) | -10.4160 | 5.3262 | -1.956 | 0.0505 |
| Education | -0.2367 | 0.1184 | -1.999 | 0.0456 |
| CT_or_MRI (Yes) | -1.6464 | 0.8532 | -1.930 | 0.0536 |
| WM-ePVS | 3.5513 | 1.7045 | 2.084 | **0.0372\*** |
| ALPS | 9.3073 | 3.8718 | 2.404 | **0.0162\*** |

## Discussion

To our knowledge, this study represents the first longitudinal investigation of changes in glymphatic imaging markers following clinical TBI. We focused on two promising and potentially widely available markers: the DTI-ALPS index and ePVS burden. Our key findings are as follows: 1) White matter ePVS burden and ALPS index are significantly correlated with age, as expected. 2) High white matter ePVS burden measured acutely following TBI is significantly associated with a higher number of post-concussion symptoms, particularly memory problems. 3) The increase in the ALPS index from the acute phase (14Day) to the chronic phase (6-12Mon) in patients with mTBI correlates with improvement in sleep measures. Furthermore, incorporating WM- ePVS burden and the ALPS index from the acute visit into patients’ basic clinical information enhances the prediction of chronic memory problems.

The PVS is an anatomical compartment that surrounds brain vascular structures^33^. The CSF-filled PVS becomes visible on MRI when enlarged, indicating impaired glymphatic function ^28, 71^. Enlarged PVS has been recognized as a hallmark of TBI since 2005^72^. Recent studies have reported increased ePVS in moderate-to-severe TBI patients^73^, special operations force solders with a history of mTBI^38^, and Vietnam War veterans with a history of TBI^74^. In moderate-to-severe TBI patients, higher ePVS burden is associated with bilateral lesions and worse verbal memory^73^. In Vietnam War veterans, high white matter ePVS is associated with poor verbal memory and elevated CSF tau levels, possibly implying impaired waste clearance^74^. In another study in Iraq/Afghanistan veterans, higher white matter ePVS was found to be associated with the number of mTBI sustained in the military, poor sleep, and severity of post-concussive symptoms^37^. In our mTBI population shortly after injury, no significant differences in ePVS burden compared to the controls were observed, but higher white matter ePVS burden was associated with acute post-injury symptoms, particularly memory problems. These findings align with recent literature on ePVS post-TBI, suggesting that acute changes in the glymphatic system may lead to more severe symptoms. Chronic ePVS burden did not correlate with symptoms at 6-12 months post-injury, possibly indicating active recovery compensating for acute structural glymphatic changes. Additionally, pre-existing conditions may contribute to increased ePVS ^33, 75^ in mTBI patients, predisposing them to higher post-concussion symptoms and memory problems.

Longitudinal changes in the white matter ePVS were not observed in the overall mTBI population or among patients with or without acute memory problems. However, the basal ganglia ePVS burden significantly increased from acute to chronic stages, although no other associations with symptoms were found, either cross-sectionally or longitudinally. The basal ganglia region is commonly associated with ePVS ^75^, and increased basal ganglia ePVS has been linked to cerebral small vessel diseases in gait disturbance^76^, and cognitive decline in early-stage Parkinson’s Disease^77^. This suggests vulnerability to vascular and glymphatic changes in this region. The stable nature of white matter ePVS over time aligns with previous reports indicating temporal stability over long follow-up periods (∼36months) ^75^. Challenges in automatic ePVS segmentation in the basal ganglia, due to reduced T1 weighted signal contrast, may contribute to variability and explain the lack of correlations with age and symptoms.

Previous TBI studies have similarly reported limited findings regarding basal ganglia _ePVS_^36,74^.

The ALPS index measures diffusion activity specifically along the medullary veins^30^, providing a sample of glymphatic activity in the brain. Despite only representing a fraction of overall glymphatic diffusion, DTI-ALPS has gained popularity, particularly due to the widespread availability of diffusion data in MRI of TBI. Reports of ALPS changes post-TBI primarily stem from retrospective studies, mostly showing reduced ALPS index in TBI patients compared to controls^50–52^. Lower ALPS index correlates with subarachnoid hemorrhage^50^, verbal memory, attention and executive functions^50, 76^, and high blood levels of neurofilament light (NfL) chain^51^, a typical TBI marker. However, Dai et al. observed increased ALPS in a group of 161 mTBI patients at an acute post-injury time (average of 5.6 days, mostly within 14 days)^78^, suggesting dynamic glymphatic activity changes post-TBI. In our study, no significant ALPS differences between control and mTBI groups were found at acute or chronic time points. However, longitudinal changes in the ALPS indexes were significantly associated with concurrent changes in sleep quality over time. Improvement in the sleep conditions correlated with increased glymphatic diffusion activity, indicating a potentially tight connection between sleep and glymphatic function, ^43, 60, 79–81^. These findings underscore the potential of ALPS index as a promising glymphatic imaging marker, linked to changes in sleep conditions following TBI, with sleep possibly mediating TBI outcomes^82–85^.

Significant age-related correlations were observed for both white matter ePVS burden and ALPS index, indicating higher ePVS burden and lower ALPS index with older age across both control and mTBI groups. While age-related decline in glymphatic function is well documented^19, 44, 86^, our study did not reveal differences in age-related slopes for these glymphatic imaging markers between the control and mTBI groups.

Despite TBI being associated with accelerated aging effects, our findings suggest that this effect may not manifest within the early chronic phase post-TBI (6-12 months), indicating a potentially longer timeframe for accelerated aging to develop.

Given that both are considered glymphatic imaging markers, a significant correlation between the ALPS index and white matter ePVS burden was found in the control population, with high ALPS index corresponding to low ePVS burden. However, this relationship was not evident in mTBI patients at either time point. Additionally, basal ganglia ePVS did not correlate with either white matter ePVS or ALPS. When incorporating glymphatic imaging markers into prediction models for chronic memory problems, acute white matter ePVS and ALPS emerged as significant variables, alongside patient education levels and the presence of CT or MRI findings. Integration of these acute glymphatic imaging markers, derived from clinically available sequences (T1w, T2w, and DTI), may significantly enhance prediction models, suggesting a potential impact of acute glymphatic function on TBI recovery. These findings highlight ePVS burden and ALPS as unique measures of glymphatic function, serving as complementary imaging markers to assess both structural and functional activity aspects of the glymphatic system.

Practical considerations regarding glymphatic imaging markers include ePVS quantification and ALPS estimation. ePVS evaluation traditionally relied on qualitative assessment, limiting accuracy and scalability. Recent studies have adopted automatic segmentation algorithms for larger datasets^29, 42, 87, 88^, emphasizing the critical importance of image quality for successful segmentation. Motion artifacts and low signal-to-noise ratio can degrade segmentation quality, leading to potential overestimation of ePVS burden. Manual inspection is essential to ensure data quality. Despite the challenges, ePVS burden remains a robust and stable parameter. Rigorous DTI preprocessing, such as eddy current correction and denoising, improves ALPS index estimation.

However, the relationship between sleep cycles and glymphatic activity may contribute to ALPS variability throughout the day, making MRI timing crucial in retrospective designs. Future studies should explore ALPS variability during different times of the day and consider MRI timing in their prospective designs.

Our study has several limitations. First, the control group included both orthopedic patients and healthy controls, potentially introduce trauma-related confounds. While we found no significant differences in glymphatic markers between these groups, trauma-related factors could still influence the results. Second, we did not observe significant symptom improvement in mTBI patients, possibly due to a selection bias favoring those with persistent symptoms. This finding aligns with larger studies like the TRACK-TBI study ^10^, suggesting that persistent symptoms at one-year post-injury are common. Our study relied on self-reported symptoms, so an important next step is to use more objective memory tests. Third, the relatively small sample size highlights the need for larger studies to better investigate longitudinal glymphatic changes and their role in TBI outcomes. Recruitment challenges, such as MRI compatibility issues with orthopedic injuries, made it difficult to exclusively use orthopedic controls for acute imaging, leading us to supplement with healthy controls. Despite these efforts, other trauma-related factors might still affect our findings. Future research should aim to include a larger and more diverse TBI population to elucidate the role of glymphatic function more accurately in TBI recovery.

## Conclusion

In summary, our longitudinal study using DTI-ALPS and ePVS burden markers reveals significant correlations with age, and high post-TBI ePVS burden is linked to increased symptoms, especially memory problems. An increase in the ALPS index correlates with improved sleep from the acute to chronic stages. Combining these markers with clinical data improves the prediction of chronic memory problems, highlighting their distinct roles in assessing glymphatic structure and activity. Early assessment of glymphatic function could be crucial for understanding TBI recovery and developing targeted interventions to improve patient outcomes.

## Data Availability

All data produced in the present study are available upon reasonable request to the authors

## Acknowledgments

The study was conducted at the University of Maryland School of Medicine Center for Innovative Biomedical Resources, Translational Research in Imaging @ Maryland (CTRIM) – Baltimore, Maryland. We thank Dr. Sepehrband for providing us with the PVS quantification tools.

